# Public health impact of catch-up vaccination or additional booster doses with pre-erythrocytic malaria vaccine R21/Matrix-M: a modelling study

**DOI:** 10.1101/2025.05.01.25326809

**Authors:** Kelly McCain, Hillary M Topazian, Joseph D. Challenger, Lucy Okell, Peter Winskill, Azra C. Ghani

## Abstract

**Background:** The pre-erythrocytic malaria vaccine R21/Matrix-M is recommended for young children, who often bear the highest burden, in malaria endemic regions. However, the small proportion of the vaccine-eligible population and waning vaccine efficacy means that routine administration is unlikely to prevent all severe cases in older children who still experience significant disease burden and mortality. Given the anticipated wide availability of R21/Matrix-M, vaccinating older age groups may be warranted.

**Methods and Findings:** Using a stochastic, individual-based transmission model of *P. falciparum* malaria, we estimate the impact of (1) a one-off catch-up campaign expanding the R21/Matrix-M vaccine-eligible population to a range of previously unvaccinated age groups between 6 months and 14 years, and (2) extra booster doses at 2, 5 and/or 10 years after the primary series in a range of transmission settings. We assume that the vaccine efficacy in older children is the same as in the standard target age group of 5–17-month-olds.

Prioritising expansion of the vaccine-eligible population through either catch-up campaigns to children under 5 years or via extra booster doses in areas of moderate or high transmission is predicted to prevent more clinical and severe cases per dose in nearly all scenarios than prioritising routine vaccination in areas of low transmission. For example, boosters 5 years post primary series averted 9 (95% CrI 5-13) severe cases per 1000 doses in a perennial setting with 45% *Pf*PR_2-10_ versus 5 (95% CrI 2-8) severe cases per 1000 doses in children receiving routine vaccination at 5% *Pf*PR_2-10_.

**Conclusion:** Additional doses of R21/Matrix-M delivered through catch-up campaigns or via extra booster doses can provide additional benefits to routine administration, but the value varies by transmission and seasonality setting. Further empirical studies, especially on vaccine efficacy in older children, are warranted to inform future policy guidance for malaria vaccination implementation.

## Introduction

Malaria continues to be a major contributor to morbidity and mortality globally, with an estimated 263 million cases in 2023 of which 94% of cases and 95% of deaths were in sub-Saharan Africa (SSA)(1). Two malaria vaccines, RTS,S/AS01 and R21/Matrix-M, have been recommended by the World Health Organization (WHO) for use in young children in SSA in 2021 and 2023, respectively, prioritising areas with moderate to high malaria transmission (2,3). Both vaccines induce anti-circumsporozoite protein (CSP) antibodies, which act during the pre-erythrocytic stage of infection when sporozoites are inoculated from an infectious bite from an *Anopheles* mosquito.

The Phase III trial of RTS,S/AS01 evaluated the vaccine efficacy in children in two age-groups – infants (6-12 weeks) and young children (5-17 months). Children received 3 doses monthly, and an additional trial arm tested the efficacy of a fourth dose 12 months post dose 3 (which we refer to as age-based implementation). Vaccine efficacy against clinical malaria over 4 years of follow-up was highest in children aged 5-36 months who received the additional fourth dose (36.3%, 95% CI 31.8-40.5%) (4). Subsequently, an alternative dosing schedule (seasonal implementation) was evaluated alongside seasonal malaria chemoprevention (SMC) in an area with intense seasonal transmission, with the first 3 doses delivered before the high transmission season and the fourth dose 12 months post dose 3 (referred to as seasonal implementation). Vaccine efficacy against clinical malaria over 5 years of RTS,S+SMC compared to SMC alone was 57.7% (95% CI 53.3-61.7) (5). The Phase III trial for R21/Matrix-M therefore evaluated vaccination of children aged 5-36 months under both age-based and seasonal implementation. Vaccine efficacy against clinical malaria after one year was 75% (95% CI 71-78) and 67% (95% CI 59-73) with seasonal and age-based implementation, respectively (6). Both efficacy against clinical malaria and antibody titres after 12 months were higher in children aged 5-17 months at the time of vaccination compared to children aged 17-36 months (6). Accordingly, WHO currently recommends a four-dose schedule delivered either through age-based or seasonal implementation, with the primary series of 3 doses given monthly to children beginning from five months of age, followed by a fourth dose 1 year post dose 3 (2). A fifth booster one year after the 4^th^ dose may be considered in seasonal settings.

Protection from both vaccines is likely to wane over time. Vaccine efficacy against clinical malaria among children aged 5-17 months in the Phase III trial for RTS,S/AS01 was 50.4% (95% CI 45.8-54.6) after 14 months (7), compared to 36.3% (95% CI 31.8-40.5) over 4 years of follow up (4). Longer-term follow-up data for R21/Matrix-M is not yet available, although efficacy was maintained for 6 months following the fourth dose (8). After protection from an imperfect intervention wanes, rebound or delayed malaria cases among the previously protected population are often observed due to a slower acquisition of natural immunity compared to an unprotected population (9). A booster dose of RTS,S/AS01 at 18 months post primary series of RTS,S/AS01 could delay a rebound in clinical malaria for up to 3 years post-booster (10). Delayed malaria may result in negative vaccine efficacy at older ages as the vaccinated cohort with waning vaccine-induced immunity and low natural immunity experiences more cases of malaria relative to an unvaccinated cohort with high levels of natural immunity (11). Over 7 years of follow-up of the RTS,S/AS01 vaccine trial in 2 sites, vaccine efficacy waned to 4.4% (−14.5-24.6) and even showed negative efficacy among highly exposed children in the fifth year post vaccination (11). Negative vaccine efficacy against severe malaria of RTS,S/AS01 in a four-dose group was also seen at the end of the seven-year follow-up period (12). Additional booster doses given before expected delayed malaria cases may prevent increased incidence of clinical malaria once vaccine-induced immunity wanes.

Globally, most of the malaria burden in moderate to high transmission settings is in children under 5 years old; however, 20% to 40% of clinical cases of malaria are in school-aged children (aged 5-15 years), with the burden of cases in this age group peaking in moderate transmission sites (13). Although school-aged children have a lower risk of symptomatic malaria infection compared to children under 5, they contribute disproportionately to the infectious reservoir as they are not typically targeted for seasonal malaria chemoprevention (SMC), sleep less frequently under bed nets, and are often the age group with the highest parasite prevalence (14–17). In low transmission settings, the risk of severe malaria is similar across all age groups (18). Furthermore, the small proportion of the current vaccine-eligible population combined with waning efficacy from the vaccine means that routine administration to young children is unlikely to generate indirect protection (i.e. herd immunity) for older children and adults (19). Therefore, expanding vaccination to older age groups to provide both direct and indirect protection may be warranted. A catch-up vaccination campaign targeting older age groups to either catch those who were missed in vaccination, as happened in Kenya during the RTS,S/AS01 pilot (20), or to expand the population of those vaccinated may also reduce transmission in the wider population.

Mathematical modelling has previously been used to estimate the public health impact of malaria vaccine introduction across SSA for both the RTS,S/AS01 and R21/Matrix-M vaccines (21–23). Here we extend the work presented in Schmit et al. for the R21/Matrix-M vaccine (22) to estimate the benefit in terms of clinical and severe cases averted of extending vaccination through catch-up campaigns to children up to age 14 years and/or via additional boosting to older children at combinations of 2, 5, and/or 10 years post primary series. Results from this study can inform ongoing discussions about wider use cases for both malaria vaccines.

## Methods

### Modelling

We used a stochastic individual-based model of *P. falciparum* malaria developed at Imperial College London (24–26) performed using the open-source *malariasimulation* R package (v 1.6.1) (27) to estimate the impact of 1) a catch-up campaign expanding the population eligible for initial R21/Matrix-M vaccination to a range of age groups between 6 months and 14 years and 2) adding extra boosters at 2, 5 and/or 10 years after the primary series to routine age-based vaccination. The model incorporates transmission dynamics between the mosquito vectors and the human hosts and a range of malaria control interventions, including pre-erythrocytic vaccines. A detailed description of the model can be found in the Supplementary Information and is summarised in brief below.

After age-dependent exposure to an infected bite, individuals become infected and, following a latent period, develop clinical disease or asymptomatic infection. A proportion of those with clinical disease are successfully treated. Treated individuals recover at a specified rate and return to the susceptible state, while those with unsuccessful treatment or no treatment move first to the asymptomatic state, then to a subpatent infection state as the parasite load falls below the microscopy-detectable limit before the individual returns to the susceptible state. The model incorporates maternally acquired immunity against clinical disease, pre-erythrocytic immunity, and blood stage immunity, each modelled to increase as a function of age and prior exposure to malaria infection. The mosquito vectors are modelled using a compartmental structure, following the female mosquito through its life stages (egg, larval, pupal, adult) with infection of adult females occurring from biting an infected human.

Anti-CSP antibodies have been shown to correlate with clinical protection against malaria (28). Using immunogenicity and case data from the Phase II R21/Matrix-M trial, modelled anti-CSP antibody titres and the relationship of antibody titre with clinical protection were fit to immunogenicity and case data from the Phase II trial for R21/Matrix-M (22,29). We modelled the R21/Matrix-M vaccine in *malariasimulation* using the median values of the derived antibody titre and vaccine efficacy parameters (Table S2) (22). Vaccine efficacy in the model is assumed to be zero until after the third dose. Antibody dynamics are described by a biphasic exponential decay model that allows short- and long-term decays of anti-CSP antibodies.

The results presented in the main text assume the same immunogenicity and vaccine efficacy regardless of age at vaccination, whilst in the Supplementary Information (p. 39 to 43), we show a sensitivity analysis assuming the peak antibody titre for age at vaccination of 5 years or older has a geometric mean ratio of 0.64 compared to the parameter fitted to data on young children, based on a study comparing the immunogenicity of RTS,S/AS02, a pre-erythrocytic vaccine similar to R21/Matrix-M, in children aged 6 to 11 years and 1 to 5 years (30). Under both assumptions, we assume that the relationship between antibody titre and vaccine efficacy remains the same regardless of age group.

### Model scenarios

We assumed that baseline prevalence of *P. falciparum* in children aged 2-10 years (*Pf*PR_2-10_) at the beginning of the simulation incorporated existing usage of malaria control interventions (such as insecticide-treated bed nets, seasonal malaria chemoprevention, indoor residual spraying), so these interventions were not modelled explicitly. Scenarios were modelled for seasonal and perennial transmission settings (Table S3) across a range of parasite prevalence (via microscopy) in children aged 2 to 10 years (*Pf*PR_2-10_ 1%-65%). Each additional vaccine intervention (expanded age groups or additional booster doses) was supplemental to routine age-based vaccination. We did not explicitly model the impact of expanded age groups or booster doses added to routine seasonal vaccination strategies as the overall impact of different delivery strategies for R21/Matrix-M in our model is similar (Fig S1) and none of the countries currently implementing pre-erythrocytic vaccination have chosen to use a seasonal routine implementation strategy (31). We modelled a population of 300,000 individuals of all ages and fixed treatment coverage with artemether-lumefantrine to 45% among clinical malaria cases (Table S3) (21). Simulations were run with a burn-in period of 20 years, following which the vaccine was introduced. Routine vaccination was continually implemented from this time onwards whilst catch-up campaigns were assumed to be delivered at a single timestep (see below). To capture the impact of extra booster doses given 10 years post primary series and any effects of delayed malaria, the simulations were run for 30 years post initial vaccination.

Vaccination strategies are summarised in Table 1. Age-based vaccination was implemented with the primary series of 3 doses starting at 6 months of age and a booster dose at 12 months post-primary series, resulting in vaccine doses at 6, 7, 8 and 20 months of age. We also evaluated one, two, or three additional booster doses at combinations of 2, 5, and/or 10 years after the primary series. Catch-up vaccination to a range of age groups was modelled as a single targeted campaign of the primary series and the booster dose 12 months later and was supplementary to ongoing routine age-based vaccination. The catch-up vaccination campaigns began in the year of vaccine introduction in perennial settings and at 5.5 months before peak clinical incidence in the year of vaccine introduction in seasonal settings. Vaccination coverage for all scenarios was assumed to be 80% for the primary series, and 80% of those who received the primary series were assumed to receive any following booster dose (64% of the target population), similar to coverage achieved during the Malaria Vaccine Implementation Programme (3) (Table S2).

**Table 1.** Summary of modelled vaccination strategies. All children receiving the vaccine through a catch-up campaign were previously unvaccinated against malaria. Age groups are inclusive (e.g. a catch-up campaign to children 6 months to 2 years old would include children up to 3 years minus 1 day old).

| **Additional vaccine strategy** | **Background vaccination** | **Additional boosters** | **Age groups targeted for catch-up campaign (inclusive)** |
| --- | --- | --- | --- |
| Routine age-based only | Primary series of three doses + single booster 12m post dose 3 to children from 6 months of age | - | - |
| Catch-up |  | - | 5 – 9 years<br>5 – 14 years<br>6 months – 2 years<br>6 months – 4 years<br>6 months – 9 years<br>6 months – 14 years |
| Additional boosters |  | 2, 5, 10,<br>2 & 5, 2 & 10, 5 & 10,<br>or 2, 5, & 10 years<br>post primary series | - |
| Catch-up + additional boosters |  | 2, 5, 10,<br>2 & 5, 2 & 10, 5 & 10,<br>or 2, 5, & 10 years<br>post primary series | 5 – 9 years<br>5 – 14 years<br>6 months – 2 years<br>6 months – 4 years<br>6 months – 9 years<br>6 months – 14 years |

Primary model outcomes were clinical and severe cases averted per 1000 people and per 1000 doses delivered compared to a counterfactual of no vaccination. Clinical and severe cases were summed over the 30 years post-introduction of vaccination. Estimates are represented as the median, and 2.5% and 97.5% percentiles of 50 parameter draws of the posterior distributions of the main malaria transmission model for each scenario.

#### Cohort analysis

We followed cohorts of children every 6 months according to the time of vaccination who were either unvaccinated or vaccinated with or without additional booster doses until the end of the 30-year simulation. For example, a cohort of children who were vaccinated in the first six months of the simulation were followed for 30 years, while children vaccinated in the first 6 months of the 25^th^ year were followed for 5 years. Cases averted, severe cases averted, doses delivered, and populations were averaged by age group across the 60 cohorts vaccinated under the routine age-based strategy. Outcomes averted were calculated by taking the difference in the number of clinical and severe cases each year per age group between the unvaccinated cohort and a vaccinated cohort in the same half year. We calculated the median of clinical and severe cases averted per year over 50 stochastic draws of the posterior distributions of the main malaria transmission model for each scenario.

#### Efficiency frontier

An efficiency frontier compares the benefits of an intervention against its cost (32). We used the number of R21/Matrix-M doses delivered as a proxy for cost and total clinical or severe cases averted as proxies for benefit compared to a no vaccination scenario, assuming a proportional increase in benefit. We plotted the median cumulative number of clinical and severe cases of malaria averted for each transmission and seasonality setting against the total number of doses delivered over the 30-year simulation under each vaccination strategy. Dominated scenarios with more doses delivered and fewer cases averted were removed from plots.

## Results

### Catch-up strategies

Fig 1 shows the modelled cases averted per dose of an additional one-off catch-up campaign delivered at vaccine introduction to different age groups compared to age-based routine implementation in a perennial setting. At moderate to high transmission (25% - 65% *Pf*PR_2-10_), the median number of cases averted per dose is highest under the currently recommended age-based schedule. In these settings catch-up campaigns to older age groups are projected to prevent similar or higher numbers of additional cases per person (Fig S2, Table 2) and to prevent substantial numbers of additional cases per dose, though the uncertainty intervals are wide (Fig 1, Table 2). The predicted efficiency of catch-up vaccination at the highest levels of transmission is reduced but is still considerable as older children are targeted, because much of the burden lies in the youngest children who would be vaccinated continuously as they age into eligibility under the routine age-based strategy (Fig 1, Fig S2).

**Fig 1.**
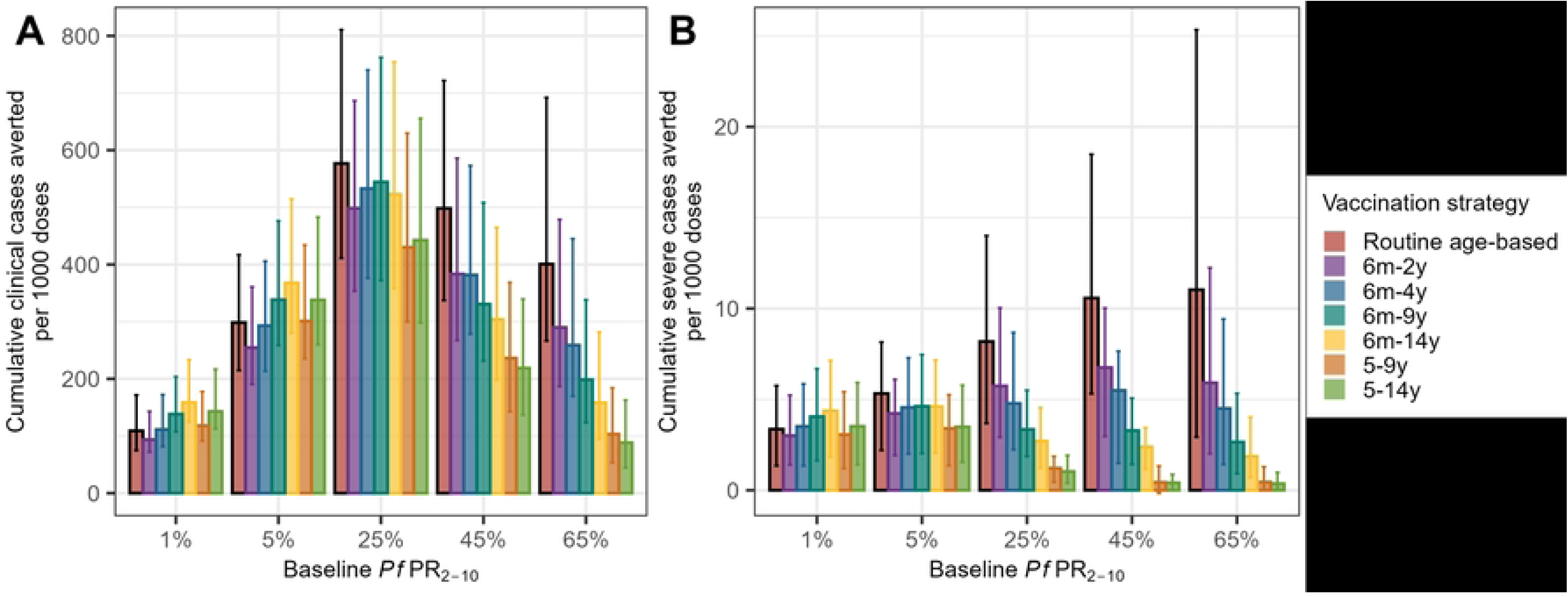
Catch-up campaign impact per 1000 doses, perennial setting. Cumulative clinical (A) and severe (B) cases averted per 1000 doses delivered to cohorts over the life-course in a 30-year simulation in a perennial setting. Routine age-based vaccination to 6-month-olds as well as catch-up vaccination in older children is assumed in all catch-up scenarios. The number of doses is calculated as the total number of doses delivered for a given cohort under the specified vaccination strategy; for example, the number of doses for a strategy “6m-2y” counts doses delivered under the catch-up strategy to children aged 6 months to 2 years alongside the routine age-based doses delivered to that cohort. The bars show median values, and the error bars show 95% credible intervals from 50 stochastic model runs. Note that the y-axes in plots A and B are different.

**Table 2.** Outcomes averted per 1000 people, per 1000 doses, and per 1000 fully vaccinated children (FVC) in a perennial setting. Strategies included in the table are either catch-up vaccination or additional boosters and are summarised over the total population and a 30-year time horizon. FVC is defined as having received the full primary series. Each of the strategies is supplementary to routine age-based vaccination that includes a single booster dose at 12 months post-primary series. The age groups listed are those targeted for a catch-up vaccination campaign (e.g. 6m-14y refers to a catch-up vaccination campaign to children between 6 months and 14 years at vaccine introduction that is supplementary to continuous routine age-based vaccination). Booster dose timing (e.g. 10y booster) is noted as the timing of additional booster dose(s) (e.g. 10y booster refers to routine age-based vaccination with a single additional booster at 10 years post primary series, so this strategy has two booster doses, the standard one at 12 months plus the additional one at 10 years). The table is grouped by transmission intensity and sorted by cases averted per 1000 doses. Median values with 95% credible intervals of 50 stochastic model runs are presented.

| Strategy | Cases averted per 1000 population | Severe cases averted per 1000 population | Cases averted per 1000 doses | Severe cases averted per 1000 doses | Cases averted per 1000 FVC | Severe cases averted per 1000 FVC |
| --- | --- | --- | --- | --- | --- | --- |
| **Low transmission: $PfPR_{2-10} = 5\%$** |  |  |  |  |  |  |
| 5y, 10y boosters | 1041 (756, 1452) | 20 (9, 32) | 293 (213, 409) | 6 (3, 9) | 1324 (960, 1846) | 25 (12, 40) |
| 10y booster | 928 (681, 1302) | 18 (8, 27) | 287 (211, 403) | 6 (2, 8) | 1179 (865, 1656) | 23 (10, 35) |
| 5y booster | 906 (646, 1273) | 18 (7, 27) | 272 (194, 382) | 5 (2, 8) | 1151 (821, 1619) | 23 (9, 35) |
| 6m-14y | 1141 (843, 1587) | 19 (8, 29) | 272 (201, 379) | 5 (2, 7) | 1027 (759, 1430) | 17 (7, 26) |
| 5-14y | 1007 (745, 1436) | 18 (8, 26) | 268 (198, 381) | 5 (2, 7) | 1010 (746, 1436) | 18 (8, 26) |
| 6m-9y | 1008 (739, 1427) | 19 (8, 28) | 264 (193, 374) | 5 (2, 7) | 998 (730, 1411) | 18 (8, 27) |
| 5-9y | 873 (643, 1249) | 17 (7, 25) | 258 (190, 368) | 5 (2, 7) | 972 (715, 1389) | 19 (8, 28) |
| 2y, 10y boosters | 936 (673, 1285) | 18 (8, 27) | 258 (185, 355) | 5 (2, 8) | 1188 (854, 1634) | 23 (10, 35) |
| 2y, 5y, 10y boosters | 1002 (731, 1406) | 20 (8, 30) | 258 (189, 362) | 5 (2, 8) | 1274 (930, 1786) | 25 (10, 38) |
| 6m-4y | 864 (623, 1234) | 16 (7, 25) | 255 (184, 365) | 5 (2, 7) | 961 (694, 1375) | 18 (8, 28) |
| 6m-2y | 803 (578, 1130) | 16 (7, 24) | 250 (180, 352) | 5 (2, 8) | 942 (679, 1328) | 19 (8, 29) |
| Routine age-based | 733 (523, 1022) | 15 (6, 22) | 247 (177, 345) | 5 (2, 8) | 932 (665, 1299) | 19 (8, 29) |
| 2y, 5y boosters | 890 (643, 1264) | 18 (8, 26) | 240 (173, 341) | 5 (2, 7) | 1132 (817, 1609) | 23 (10, 33) |
| 2y booster | 762 (542, 1083) | 15 (7, 23) | 224 (159, 318) | 5 (2, 7) | 969 (688, 1378) | 20 (8, 30) |
| **Moderate transmission: $PfPR_{2-10} = 25\%$** |  |  |  |  |  |  |
| 5y, 10y boosters | 2334 (1713, 3100) | 28 (16, 42) | 660 (484, 874) | 8 (5, 12) | 2976 (2182, 3948) | 36 (21, 53) |
| 10y booster | 2015 (1492, 2677) | 25 (15, 39) | 625 (462, 829) | 8 (5, 12) | 2568 (1899, 3404) | 32 (19, 49) |
| 5y booster | 2043 (1530, 2761) | 27 (16, 40) | 613 (458, 830) | 8 (5, 12) | 2599 (1945, 3520) | 34 (21, 51) |
| 2y, 5y, 10y boosters | 2271 (1655, 3087) | 29 (17, 43) | 587 (428, 799) | 7 (4, 11) | 2893 (2110, 3933) | 37 (21, 55) |
| Routine age-based | 1697 (1261, 2305) | 25 (14, 38) | 573 (426, 779) | 8 (5, 13) | 2160 (1606, 2937) | 32 (18, 48) |
| 2y, 10y boosters | 2066 (1554, 2785) | 28 (15, 40) | 572 (429, 772) | 8 (4, 11) | 2633 (1978, 3555) | 35 (20, 51) |
| 6m-2y | 1799 (1340, 2487) | 26 (15, 39) | 562 (418, 776) | 8 (5, 12) | 2120 (1577, 2925) | 31 (18, 46) |
| 2y, 5y boosters | 2069 (1515, 2804) | 28 (16, 41) | 559 (410, 759) | 7 (4, 11) | 2635 (1931, 3574) | 35 (20, 53) |
| 6m-4y | 1883 (1404, 2562) | 27 (15, 40) | 556 (415, 756) | 8 (5, 12) | 2097 (1566, 2849) | 30 (17, 45) |
| 5-9y | 1840 (1371, 2535) | 25 (15, 39) | 544 (405, 748) | 7 (4, 11) | 2052 (1526, 2821) | 28 (16, 43) |
| 6m-9y | 2056 (1540, 2819) | 27 (15, 41) | 540 (405, 740) | 7 (4, 11) | 2039 (1527, 2791) | 27 (15, 41) |
| 5-14y | 2000 (1465, 2699) | 25 (15, 39) | 532 (390, 717) | 7 (4, 10) | 2006 (1471, 2703) | 25 (15, 39) |
| 6m-14y | 2179 (1595, 2979) | 27 (15, 41) | 521 (381, 711) | 6 (4, 10) | 1965 (1439, 2683) | 24 (14, 37) |
| 2y booster | 1743 (1302, 2404) | 26 (14, 38) | 512 (382, 707) | 8 (4, 11) | 2221 (1659, 3068) | 33 (17, 49) |
| **Moderately high transmission: $PfPR_{2-10} = 45\%$** |  |  |  |  |  |  |
| 5y, 10y boosters | 2498 (1803, 3447) | 29 (16, 45) | 706 (509, 973) | 8 (4, 13) | 3187 (2295, 4389) | 37 (20, 57) |
| 5y booster | 2260 (1639, 3087) | 28 (16, 45) | 678 (493, 927) | 9 (5, 13) | 2878 (2090, 3934) | 36 (20, 57) |
| 10y booster | 2153 (1560, 2995) | 28 (14, 44) | 667 (483, 930) | 9 (4, 14) | 2740 (1985, 3819) | 36 (18, 56) |
| Routine age-based | 1934 (1393, 2605) | 29 (15, 46) | 654 (470, 878) | 10 (5, 15) | 2463 (1773, 3310) | 37 (19, 58) |
| 2y, 5y, 10y boosters | 2505 (1792, 3420) | 30 (16, 49) | 647 (463, 884) | 8 (4, 13) | 3190 (2285, 4359) | 38 (21, 62) |
| 6m-2y | 2015 (1471, 2734) | 30 (15, 47) | 631 (461, 853) | 9 (5, 15) | 2378 (1738, 3215) | 35 (18, 55) |
| 2y, 10y boosters | 2282 (1649, 3098) | 29 (16, 47) | 631 (456, 854) | 8 (4, 13) | 2906 (2101, 3937) | 37 (20, 61) |
| 2y, 5y boosters | 2304 (1674, 3127) | 30 (17, 48) | 623 (452, 843) | 8 (4, 13) | 2934 (2133, 3977) | 38 (21, 61) |
| 6m-4y | 2074 (1500, 2869) | 30 (15, 46) | 614 (443, 850) | 9 (5, 14) | 2317 (1672, 3206) | 33 (17, 52) |
| 5-9y | 2024 (1481, 2758) | 28 (15, 45) | 597 (438, 815) | 8 (4, 13) | 2253 (1652, 3074) | 31 (17, 50) |
| 2y booster | 1993 (1479, 2729) | 29 (14, 46) | 586 (434, 801) | 9 (4, 14) | 2539 (1884, 3471) | 37 (18, 59) |
| 6m-9y | 2150 (1530, 2994) | 29 (16, 47) | 565 (401, 786) | 8 (4, 12) | 2131 (1513, 2968) | 29 (16, 47) |
| 5-14y | 2067 (1500, 2811) | 28 (14, 45) | 550 (398, 745) | 7 (4, 12) | 2075 (1503, 2812) | 28 (14, 45) |
| 6m-14y | 2186 (1601, 3066) | 29 (15, 46) | 521 (383, 731) | 7 (4, 11) | 1968 (1444, 2760) | 26 (14, 42) |

In contrast, at low transmission, catch-up campaigns to older children are projected to avert more cases and similar numbers of severe cases per dose compared to delivery under the recommended age-based schedule (Fig 1). In settings with moderate to high transmission, the efficiency of catch-up campaigns in averting severe cases is lower compared to routine age-based vaccination, while efficiency remains considerable compared to age-based vaccination up to 25%-45% *Pf*PR_2-10_ for clinical cases (Fig 1).

Comparing across transmission settings, the value of catch-up campaigns per dose delivered to children under 5 years is greatest in 25% - 45% *Pf*PR_2-10_ settings when considering severe disease and in 25% *Pf*PR_2-10_ settings when considering clinical disease (Fig 1), due to the projected higher burden of delayed malaria at high transmission. For example, over the 30-year simulation, supplementary catch-up campaigns targeting children aged 6 months to 4 years are projected to avert 9 (95% CrI 5-14) severe cases per 1000 doses in a perennial setting with 45% *Pf*PR_2-10_ versus 5 (95% CrI 2-7) severe cases per 1000 doses with only routine vaccination at 5% *Pf*PR_2-10_ (Table 2). Prioritising catch-up vaccination to children up to 5 years of age in moderate transmission settings may be more efficient compared to routine vaccination in lower transmission settings when considering both clinical and severe cases.

In seasonal settings, the number of projected cases averted per dose is generally higher than in perennial settings because the catch-up campaigns were timed in our model to be prior to the peak in transmission and hence provided the highest level of protection during the period of highest risk (Fig S3). Routine age-based vaccination is projected to avert fewer clinical and severe cases per dose compared to age-based + catch-up strategies in all seasonal transmission settings, except regarding severe cases at 45% - 65% *Pf*PR_2-10_ where routine vaccination is most efficient (Fig S3). The target age ranges for catch-up vaccination that avert more cases per dose than routine-age based vaccination become narrower as transmission increases (e.g. at 5% *Pf*PR_2-10_, all catch-up campaigns are more efficient while at 65% *Pf*PR_2-10_, only catch-up campaigns to children aged 6 months to 2 years outperforms routine age-based vaccination).

Figure 2 shows how the projected number of clinical cases averted as a child ages changes depending on the age at first vaccination. In a low transmission setting (5% *Pf*PR_2-10_), the greatest number of cases averted immediately after vaccination is in the oldest ages at vaccination (here illustrated in green as 12-13 years) (Fig 2A). However, over the whole life course, first vaccination to older children is still marginally more efficient, but a similar number of cases are averted across all ages at vaccination (Fig 2B). In contrast, at high transmission (45% *Pf*PR_2-10_), first vaccination to older children results in fewer total cases averted over the life course compared to first vaccination to younger children; for example, first vaccination to children aged 0.5-1 years averted 1710 (95% CrI 1176-2644) cases per 1000 population, while vaccination to children aged 12-13 years averted 706 (95% CrI 421-1110) cases per 1000 population over the life course.

**Fig 2.**
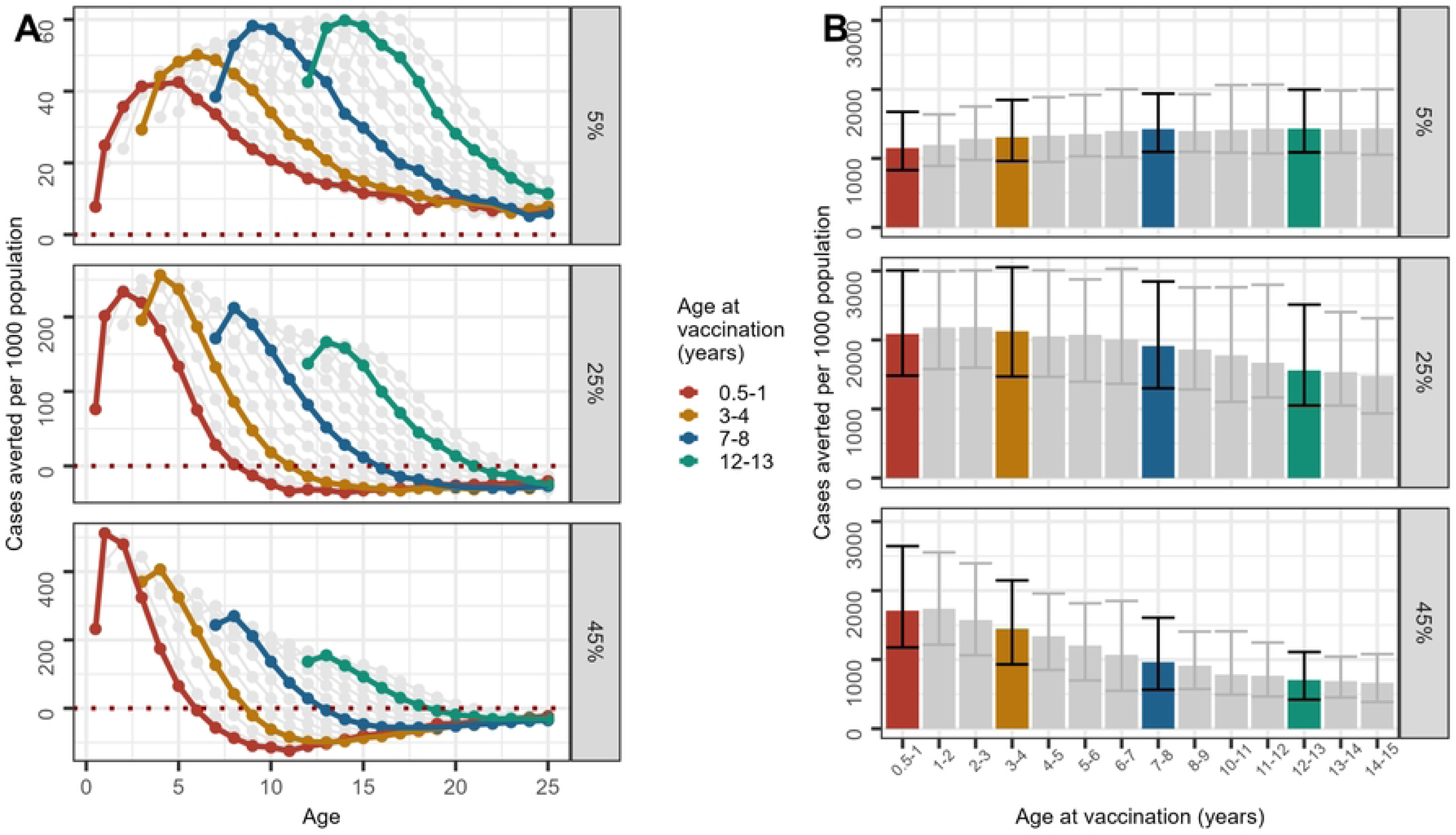
Clinical cases averted per 1000 fully vaccinated population. Cases averted per 1000 people are summarised over 30 years in a cohort of children aged 6 months to 14 years old at vaccination in a perennial setting. Panel A shows the cases averted per 1000 people by age. Each point on Panel A refers to the number of cases averted over a 6-month duration for each 6-month age group in a cohort that was vaccinated at specified ages defined by colours. Panel B shows the sum of cases averted per 1000 people over all age groups within a cohort of people vaccinated at specified ages (x-axis). All ages at vaccination (age groups from 6 months to 14 years) are plotted; a few ages at vaccination have been highlighted. The age range shown in Panel A is up to 25 years of age; totals plotted in Panel B sum cases averted in cohorts over the 30-year simulation (e.g. the cohort vaccinated at age 14 would be 44 years old at the end of the simulation).

Both routine vaccination and routine plus catch-up campaigns are projected to modify the age distribution of clinical cases (Table S4). At low perennial transmission (5% *Pf*PR_2-10_), 10% (95% CrI 6-17) of cases are estimated to be among children under 5 years prior to vaccination, reducing to 6% (95% CrI 4-10) with routine vaccination and similar values with supplemental catch-up campaigns. At high perennial transmission (45% *Pf*PR_2-10_), 41% (95% CrI 27-66) of cases are estimated to be in children under 5 years prior to vaccination, in line with other modelling studies (13). The percentage of cases in children under 5 years fell to 29% (95% CrI 19-46) with routine vaccination and to 28% (95% CrI 18-44) with additional catch-up campaigns (Table S4). This change in age distribution follows shifting age burdens observed after effective control interventions such as indoor residual spraying and the distribution of long-lasting insecticide-treated nets among children (33) or prophylaxis given to infants (34) seen in field settings and aligns with other modelling studies showing infection-blocking interventions such as pre-erythrocytic vaccination or seasonal malaria chemoprevention shifting the age distribution (35–37).

### Additional booster doses

Figure 3 shows the impact of supplemental booster doses delivered in combinations of 2, 5, and/or 10 years added to routine age-based vaccination. The scenarios with boosters at 5, 10, and 5 and 10 years provide similar or slightly higher efficiency in terms of clinical cases averted per dose compared to routine age-based vaccination alone (Fig 3A). The addition of a booster at 2 years alone or combined with other boosters at 5 and/or 10 years tends to be less efficient compared to later booster doses (Fig 3A). The median efficiency of extra booster doses for preventing severe cases compared to age-based vaccination is similar or slightly higher at the lowest transmission levels (1% and 5% *Pf*PR_2-10_), similar at 25% *Pf*PR_2-10_, and lower in settings with 45% or higher parasite prevalence (Fig 3B). A supplemental booster dose two years after the primary series averts fewer clinical cases in all settings compared to delaying the extra dose to 5 and/or 10 years, likely because of re-vaccination prior to waning of vaccine-induced protection (Fig 3A). However, in high transmission settings, a booster dose two years after the primary series is more efficient against severe cases compared to other booster dose timings (Fig 3B).

**Fig 3.**
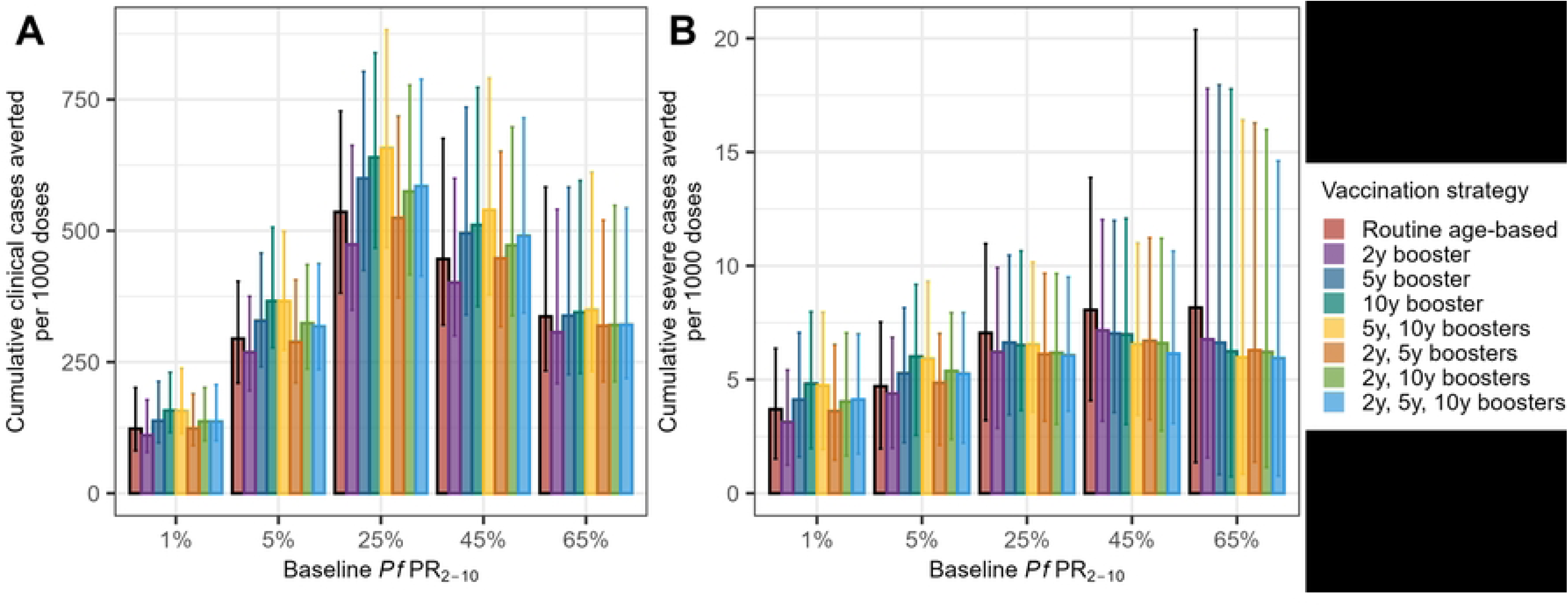
Additional booster impact per 1000 doses, perennial setting. Cumulative clinical cases (A) and severe cases (B) averted per 1000 doses in a perennial setting, by strategies where boosters were delivered at 12 months and 2 years, 5 years and/or 10 years in addition to routine vaccination at age 6 months. The bars show median values, and the error bars show 95% credible intervals of 50 stochastic model runs. Outcomes are summarised over the final 15 years of the simulation so that each scenario had 15 years of continuous vaccination to young children prior to calculating cases averted. This allows for a fairer comparison between strategies with different booster dose timing. Note that the y-axes in plots A and B are different.

Comparing across transmission settings, the per-dose impact when considering clinical cases of all age-based strategies with additional boosters increases with transmission intensity up to 45% *Pf*PR_2-10_ and is similar at 65% *Pf*PR_2-10_ even when delivering additional booster doses to older children. However, introducing extra booster doses to children in high transmission settings is predicted to be more efficient for both severe and clinical cases than implementing routine age-based vaccination in lower transmission settings (Fig 3). For example, over the entire 30-year simulation, adding an additional booster 5 years post primary series averted 9 (95% CrI 5-13) severe cases per 1000 doses in a perennial setting of 45% *Pf*PR_2-10_ versus 5 (95% CrI 2-8) severe cases per 1000 doses in children receiving routine vaccination at 5% *Pf*PR_2-10_ (Table 2).

The patterns of projected impacts of the additional booster strategies are relatively similar for both clinical cases and severe cases averted per population at low transmission where the age pattern of malaria burden is evenly distributed across age groups and where delayed malaria due to waning vaccine-induced immunity is less evident (Fig S5A). However, at higher transmission, the relative impact of the additional booster doses on clinical cases versus severe cases diverges. At the highest levels of transmission (45% and 65% *Pf*PR_2-10_) there is significant benefit in averting clinical cases but less clear benefit in averting severe disease with extra booster doses, due to the concentration of severe disease in younger age groups. In contrast, at lower transmission, additional boosters provide added protection against severe disease in the older population (Fig S5B).

In cohorts vaccinated with a routine age-based strategy (Fig 4), additional boosters recover the vaccine-induced protection that otherwise wanes over time, though the extent to which protection is recovered varies by transmission intensity. At 25% *Pf*PR_2-10_ and above, the number of cases averted by age group is projected to fall over the first few years after vaccination; additional booster doses are projected to both delay and reduce the subsequent rebound. However, at 65% *Pf*PR_2-10_, two to three additional boosters delivered after protection from the primary series wanes are not sufficient in themselves to entirely offset the rebound in incidence. Despite the rebound, the large number of cases averted in young children means that the total numbers of clinical and severe cases averted per dose and per person remain positive (Figs 1, S5). The single booster dose at 10 years post primary series results in more cases averted compared to the scenarios where booster doses were also given earlier (e.g. at 5 and 10 years, 2 and 10 years, or 2, 5, and 10 years) (Fig 4).

**Fig 4.**
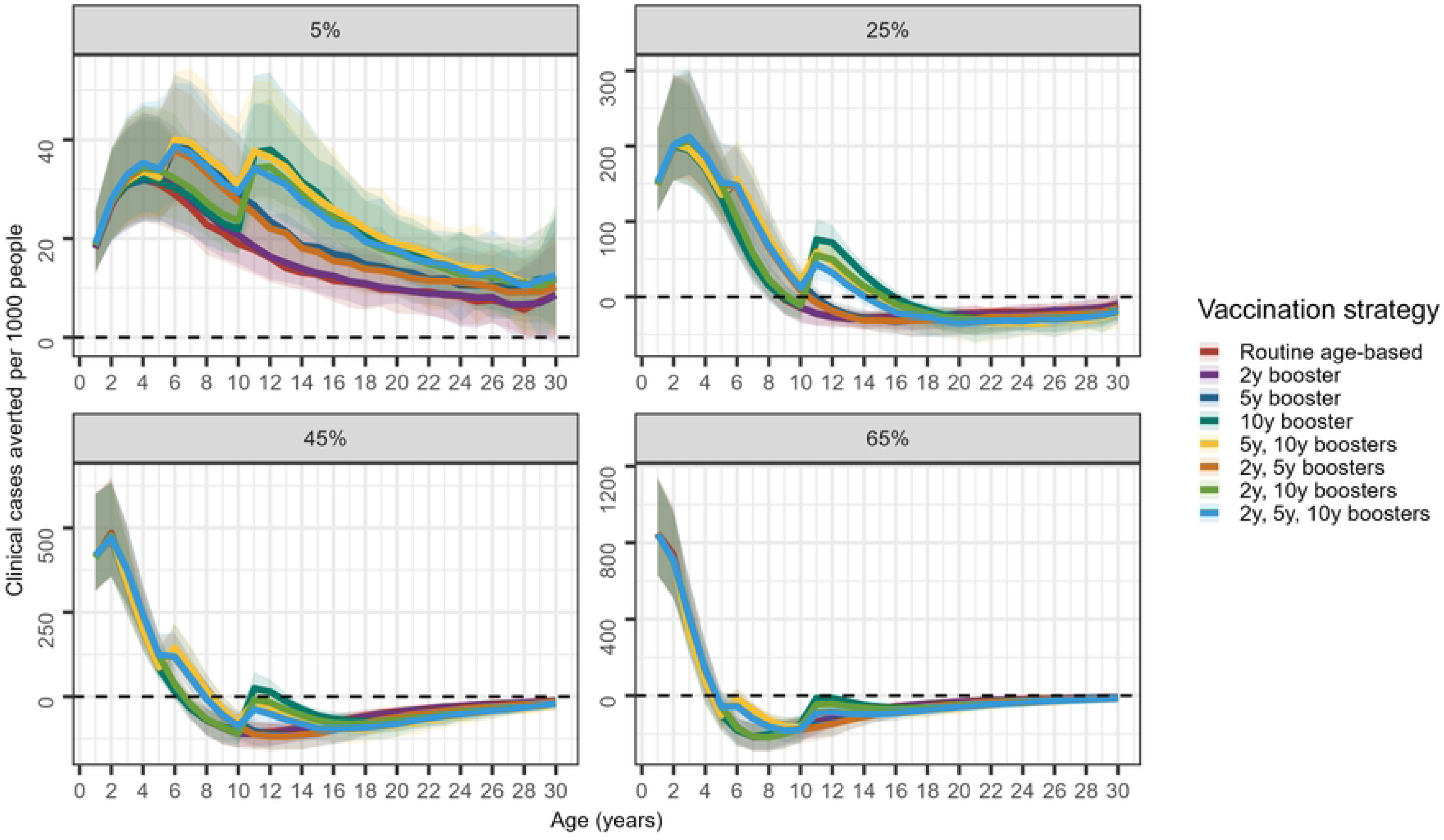
Additional booster impact: cases averted per 1000 population by age. Cohort view of clinical cases averted per 1000 people in each 1-year age group compared to a no vaccination baseline scenario in a perennial setting. For example, for age 10 this is a snapshot summary of the state of all children aged 10 at any point during the simulation across all vaccinated cohorts. The coloured lines are the median values, and the shaded regions are the 95% credible intervals over the 60 cohorts and 50 stochastic parameter draws.

In contrast to their impact on clinical cases, extra boosters provide limited additional benefit in averting severe cases to the routine age-based primary series, especially at moderate to high transmission (Fig S6). At 25%, 45% and 65% *Pf*PR_2-10_, vaccinated children who didn’t receive additional boosters are predicted to have higher incidence of clinical malaria than the unvaccinated cohort from once the vaccine protection wanes to well into adulthood, but similar incidence for severe malaria starting around 8-12 years post primary series. This is due to severe disease susceptibility being partially physiological with the peak risk at younger ages and is consistent with the 7-year follow-up data of RTS,S/AS01 (11).

### Combination of catch-up vaccination and additional boosters

At each perennial transmission intensity and seasonality, the greatest number of severe cases averted per 1000 people is projected to occur when catch-up vaccination to children aged 6 months to 14 years is combined with three extra booster doses at 5 and 10 years post primary series, though the estimates are very similar in many of the combination strategies. In perennial settings, the number of clinical cases averted per 1000 people for this strategy ranges from a median of 1444 (95% CrI 1088-1988) at 5% *Pf*PR_2-10_ to 2775 (95% CrI 1969-3838) at 45% *Pf*PR_2-10_ (Table S5).

Over the 30-year simulation, the greatest number of cases averted per 1000 doses is projected to occur with catch-up vaccination to children 5-14 years old (302 (95% CrI 226-423)), 6 months to 9 years old (302 (95% CrI 224 - 416)), and 6 months to 14 years old (302 (95% CrI 228, 416)), each combined with two extra booster doses at 5 and 10 years post primary series at 5% *Pf*PR_2-10_.The most efficient strategy is catch-up vaccination to children 6 months to 2 years combined with two extra booster doses at 5 and 10 years at 25% (641 (95% CrI 477-860)), 45% (684 (95% CrI 491-933)), and 65% *Pf*PR_2-10_ (555 (95% CrI 408-844)) (Table S5).

### Efficiency

Figure 5 shows the efficiency frontier for a range of different vaccination scenarios at different transmission levels for perennial settings (with similar results for seasonal settings shown in Fig S7). If additional doses are available for supplementation of the routine age-based strategy, the next most efficient strategy within every transmission setting is adding catch-up vaccination to children aged 6 months to 2 years. With more available doses, the next most efficient strategies comprise different combinations of expanding catch-up campaigns to children up to 14 years of age and including additional booster doses, depending on transmission intensity and seasonality. The pattern is similar for severe cases at low transmission; however, the value of additional catch-up vaccination in older age groups at moderate to high transmission is less clear than the value against clinical disease as it depends on the extent to which severe disease is experienced at older ages. The most efficient strategies in averting severe cases at high transmission are combinations of catch-up campaigns to children up to 4 years of age with extra booster doses.

**Fig 5.**
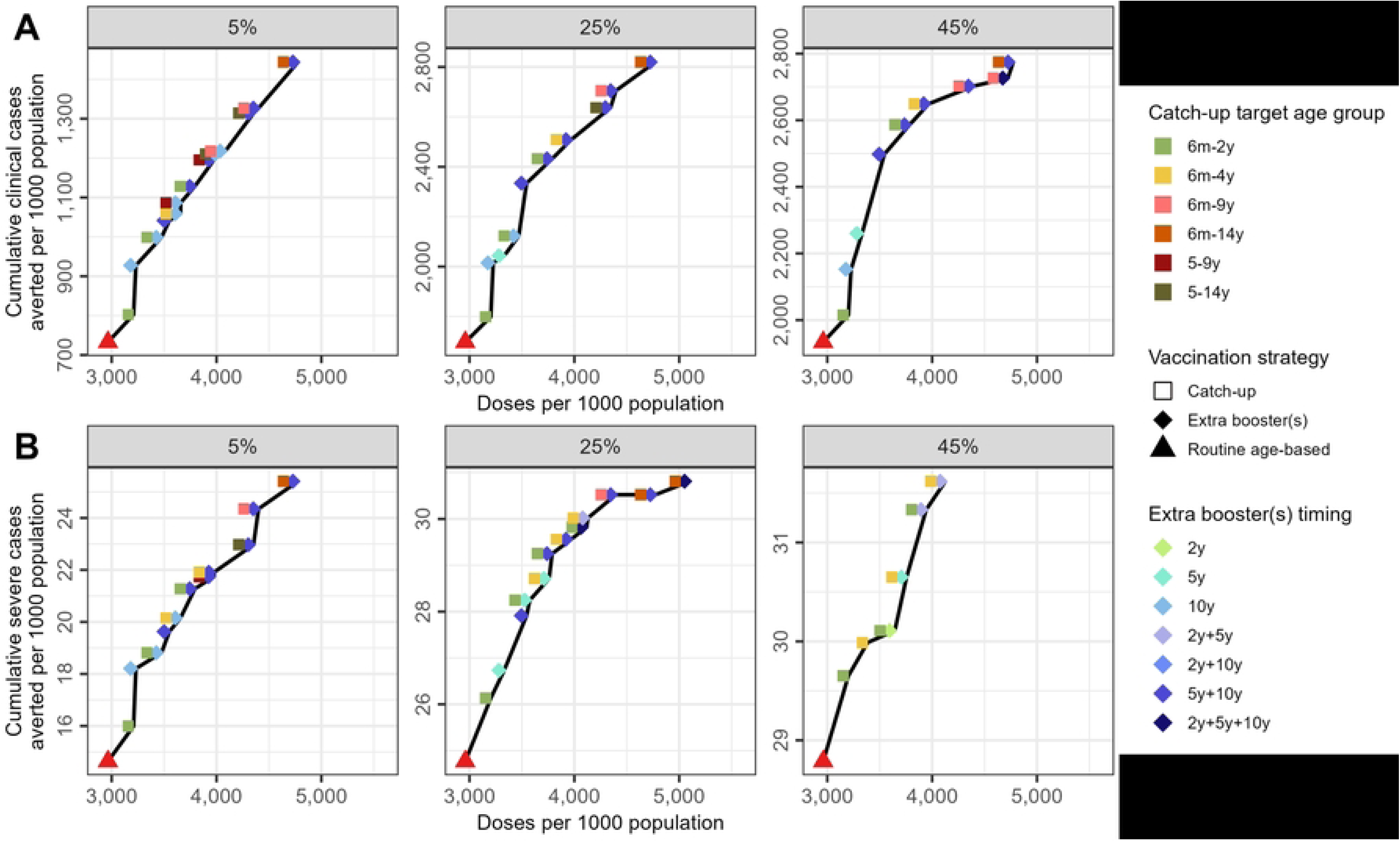
Efficiency frontier: cases averted as the number of doses available increases. Perennial settings are plotted. Circular points show vaccination strategies with catch-up campaigns supplementary to routine age-based vaccination. Triangular points show routine age-based strategies without catch-up campaigns. Labels indicate the strategy presented by the routine age-based/catch-up stratification followed by the number of booster doses (up to two additional boosters at 5 years and 10 years post routine age-based primary series). The black line connects the points lying on the efficiency frontier; all dominated strategies (higher cost, fewer cases averted) were removed. Panel A represents efficiency frontiers for clinical cases of malaria and panel B represents efficiency frontiers for severe cases of malaria in perennial settings.

### Sensitivity Analysis

The main analysis assumes that the immunogenicity and efficacy of R21/Matrix-M is the same regardless of age. In the Supplementary Information, we present the sensitivity analysis results assuming the immunogenicity is scaled by 0.64 for children aged 5 years and older as described in the Methods. As expected, the scenarios where older children are vaccinated have slightly lower impacts in terms of cases and severe cases averted, but the difference is slight (Figs S9, S10, S11, Table S7).

## Discussion

Our results confirm that the largest incremental benefit of malaria vaccine implementation is through introduction of routine age-based vaccination as currently recommended by WHO. They further demonstrate that additional doses delivered through either expansion of targeted age groups through catch-up campaigns or via extra booster doses can provide additional benefits but that the value varies by transmission and seasonality setting.

Catch-up vaccination aims to protect children who may have missed doses and/or be older than the recommended age group and is commonly implemented for routine childhood vaccinations, including Hepatitis B, and measles/mumps/rubella vaccines (38,39). We found that except at very high transmission intensity, investment in supplementary catch-up vaccination campaigns with R21/Matrix-M to children up to age two could increase the number of cases averted per person compared to routine vaccination and is the next most efficient catch-up strategy per dose to introduce following routine vaccination if additional doses are available over 30 years. Catch-up vaccination targeting children over 2 years also averts more cases per person, is relatively efficient especially for clinical cases, and may also improve equity in access to malaria control interventions. School-aged children, a population that often has high prevalence of malaria, are less likely to use ITNs and may be ineligible for SMC since the age of eligibility is typically only up to 59 months of age, are often unprotected (14–16,16,17). Catch-up vaccination to older age groups would protect these vulnerable children.

The RTS,S/AS01 vaccine was evaluated with and without booster doses in clinical trials; children who received booster doses experienced a longer period of protection (4). A fourth dose of R21/Matrix-M 12 months post primary series increased the anti-CSP antibody titres back to levels close to the peak values after the primary series and boosted efficacy (29). Given that waning protection from these vaccines has been observed, a fifth dose two years post primary series is already recommended in settings with high malaria transmission risk and in seasonal settings (2).We show that additional booster doses delivered 2, 5 and/or 10 years following dose 3 could avert more cases and be more efficient than boosting solely at 2 years following dose 3 in most settings. This is likely because the extra dose at 2 years would be delivered before full protection from the vaccine wanes, limiting its additional benefit compared to delaying it to 5 years post dose 3. However, because the rebound of the incidence of malaria occurs more rapidly in high transmission settings, providing an additional booster sooner than 5 years post routine age-based vaccination may be warranted. Timing of additional boosters could therefore be tailored based on local evidence on the timing of rebound. Our results also suggest that extra boosters would avert additional severe cases per dose only in the lowest transmission settings where the period at-risk extends to school-aged children and adults. However, given the changing epidemiology of malaria that is resulting in a shift in both clinical and severe disease to older age-groups as natural immunity is reduced, empirical data are needed to support decisions on periods of highest risk.

If there is sufficient dose supply and funding, combining both catch-up vaccination to wider age groups and extra booster doses is projected to avert more cases per person than either catch-up vaccination or additional boosters alone, and in many settings, the combination strategies were the most efficient. If extra vaccine doses are available in low transmission settings, our results suggest that it would be most advantageous to vaccinate as many people as possible, prioritising progressively older age groups. In these settings, partially protecting a larger proportion of the population is likely to result in a higher degree of indirect protection. At high transmission, our results suggest that expanding age groups via catch-up campaigns has less of an impact as older children will already have acquired natural immunity. Hence, extra doses may be more impactful when delivered as additional boosters to children who were previously vaccinated. However, with the consequential funding cuts announced in early 2025 to government foreign aid budgets and global aid programs, including the Global Fund and Gavi, which would have provided large investments in malaria vaccines, the future of malaria vaccination is uncertain (40–43).

The choice of vaccination strategy may also depend on its acceptability to the target population. Although acceptance of the RTS,S/AS01 vaccine is high (44), some implementation strategies were more successful than others. The Malaria Vaccine Implementation Programme (MVIP) for RTS,S/AS01 showed high community acceptance and demand for the vaccine, but they found that achieving high coverage of the fourth dose was challenging (3).

Our study has some limitations. First, in the absence of data across all age-groups for R21/Matrix-M, we assume that anti-CSP antibody response and subsequent vaccine efficacy are the same regardless of age. The Phase III R21 trial data showed both lower antibody titres and vaccine efficacy in the 18–36-month group compared to the 5-17-month group which was reflected in lower vaccine efficacy (6). In the Phase III RTS,S/AS01 trial, higher baseline anti-CSP antibodies prior to vaccination, indicating more exposure to malaria, were associated with higher anti-CSP antibody titres post vaccination (28,45) whilst a pooled analysis of data from Phase II RTS,S/AS01 studies showed vaccine efficacy to be highest in very young children, lower in children around 12 months of age and then increase in children up to 5 years old (46). Factors like the maturity of immune system, prior exposure to malaria, and the quality of immune response (antibody avidity) influence the immune response and vaccine efficacy; therefore, further data are needed for R21/Matrix-M to be able to capture potential variation in vaccine efficacy by age across different transmission settings.

We also made several simplifying assumptions. Firstly, we assumed a population with no importation, which may overestimate the benefits of vaccination at lower transmission where indirect protection will be more substantial. Secondly, we assumed that the catch-up vaccination campaign started and finished at the same time as vaccine introduction, which may lead to overestimation of its impact, as in reality catch-up campaigns may proceed over several years. Thirdly, whilst the efficiency frontier is a useful method to compare strategies, model uncertainty means that the most efficient scenario for a certain number of doses is also uncertain, and it was not computationally feasible to capture this uncertainty. Finally, we did not capture the incremental costs of catch-up campaigns and booster dose strategies, which could modify the efficiency frontier if translated into cost. Catch-up campaigns may be expensive because they require infrastructure outside of the Expanded Program for Immunization (EPI) (47), while routine age-based vaccination could be conducted at already-functioning EPI sites on established schedules. Additionally, the interventions with extra booster doses spread the costs of delivery and implementation over multiple years and would also require additional visits that could increase costs to the health system. Given this uncertainty, we chose not to conduct a full cost-effectiveness analysis at this stage.

In previous model predictions of the impact of R21/Matrix-M under routine vaccination over 15 years, the impact on clinical cases from routine age-based vaccination increased with transmission intensity (22), while we found that after a peak at moderate transmission, impact per dose decreased with increasing transmission over a 30-year time horizon. Our model runs were twice as long, so summary calculations included a longer period of rebound malaria, which is more pronounced at high transmission. However, the impact on deaths in Schmit et al. plateaued at high transmission, while our model runs showed a continuous increase in impact on severe cases as transmission increased.

Our model predictions showed a slightly higher benefit in seasonal compared to perennial settings for all catch-up strategies. The per-dose benefit of catch-up campaigns was lower relative to routine vaccination in seasonal settings, whereas in perennial settings, the routine strategy was often most efficient. Both differences between seasonality settings are artefacts of the optimised timing of the catch-up campaigns in seasonal settings prior to the peak in transmission.

Routine age-based vaccination with R21/Matrix-M in settings with endemic malaria delivered through an age-based or seasonal four-dose schedule was recommended by WHO in October 2023 with 15 countries planning to introduce vaccination with R21/Matrix-M in 2024 via their EPI programmes (48,49). As the production of R21/Matrix-M increases to its predicted 200 million doses per year by 2026 (50), a sum that would cover the projected demand estimated by Gavi of 80-100 million annual doses by 2030 (51), countries may wish to expand vaccination eligibility to older age groups or increase the number of booster doses administered if funding mechanisms remain viable. Our results demonstrate that expanding eligible age groups through catch-up campaigns, delivering additional boosters, or a combination of the two strategies could avert additional cases and thus provide a useful additional strategy to further reduce malaria burden. Empirical studies to evaluate the impact of both strategies are therefore warranted to inform future policy guidance regarding wider use of malaria vaccines.

## Data Availability

All relevant data are within the manuscript and its Supporting Information files. The code required to reproduce the results in this study is available at htps://github.com/kellymccain28/catchup_extraboosters. The malaria transmission model used in the study is freely available at htps://github.com/mrc-ide/malariasimulation.

https://github.com/mrc-ide/malariasimulation

https://github.com/kellymccain28/catchup_extraboosters

## Author contributions

All authors conceived the study. KM developed the model code, undertook the data analysis and modelling, and wrote the first draft of the manuscript. All authors reviewed and contributed to the final manuscript and were responsible for the decision to submit the manuscript for publication. All authors attest that they meet the ICMJE criteria for authorship.

## Data sharing

The code required to rerun the analysis in this study is available at https://github.com/kellymccain28/catchup_extraboosters. The transmission model used in the study is freely available at https://github.com/mrc-ide/malariasimulation/.

## Funding

KM acknowledges research funding from the Imperial College President’s Scholarship. All authors acknowledge funding from the MRC Centre for Global Infectious Disease Analysis (reference MR/X020258/1), funded by the UK Medical Research Council (MRC). This UK funded award is carried out in the frame of the Global Health EDCTP3 Joint Undertaking. JC acknowledges research funding from Open Philanthropy (reference GV673604959).

## Notes

### Competing Interest Statement

The authors have declared no competing interest.

### Funding Statement

The author(s) received no specific funding for this work.

